# Can Large Language Models Provide Emergency Medical Help Where There Is No Ambulance? A Comparative Study on Large Language Model Understanding of Emergency Medical Scenarios in Resource-Constrained Settings

**DOI:** 10.1101/2024.04.17.24305971

**Authors:** Paulina Boadiwaa Mensah, Nana Serwaa Quao, Sesinam Dagadu, Cohort 2, Project Genie Clinician Evaluation Group

## Abstract

The capabilities of Large Language Models (LLMs) have advanced since their popularization a few years ago. The healthcare sector operates on, and generates a large volume of data annually and thus, there is a growing focus on the applications of LLMs within this sector. There are a few medicine-oriented evaluation datasets and benchmarks for assessing the performance of various LLMs in clinical scenarios; however, there is a paucity of information on the real-world usefulness of LLMs in context-specific scenarios in resourceconstrained settings. In this study, 16 iterations of a decision support tool for medical emergencies using 4 distinct generalized LLMs were constructed, alongside a combination of 4 Prompt Engineering techniques: In-Context Learning with 5-shot prompting (5SP), chain-of-thought prompting (CoT), self-questioning prompting (SQP), and a stacking of self-questioning prompting and chain-of-thought (SQCT). In total 428 model responses were quantitatively and qualitatively evaluated by 22 clinicians familiar with the medical scenarios and background contexts. Our study highlights the benefits of In-Context Learning with few-shot prompting, and the utility of the relatively novel self-questioning prompting technique. We also demonstrate the benefits of combining various prompting techniques to elicit the best performance of LLMs in providing contextually applicable health information. We also highlight the need for continuous human expert verification in the development and deployment of LLM-based health applications, especially in use cases where context is paramount.

## 1. Introduction

In a previous study, it was observed that the outputs of generalized Large Language Models (LLMs) varied significantly when prompted to provide first aid instructions for managing medical emergencies, depending on whether background context was included or excluded. Mensah et al. (2024). For example, keeping prompt instructions, model type and model parameters constant, the addition or exclusion of background context as simple as: “*Location: busy market in rural Atebubu*, Ghana. There is a maternity home 5km *away, a chemist, 20km away, a CHPS compound 39*.*8km away, and a hospital 50km away*”, changes at least one of the following parts of the response: the possible diagnoses given, the order of the possible diagnoses given, the order of first aid instructions given, and the content of first aid instructions given. In managing medical emergencies, altering the sequence of potential diagnoses and first aid instructions, even if the content remains unchanged, can impact patient outcomes. This is because healthcare providers often address issues in the order suggested, and timely intervention is crucial for addressing the most critical problems effectively. In internal experiments, this pattern was consistent across several popular LLMs tested, with a notable difference in semantic similarity between model outputs, ranging from 20% to 30%, when background context was omitted vs included in the prompt. In building LLM-based tools for clinical applications, that loss can be clinically costly. The implication is that to derive genuine clinical utility from LLMs, it is insufficient for them to meet standard medical benchmarks; their performance in diverse contexts must be assessed. Otherwise, responses deemed beneficial in certain situations may not only prove ineffective in others but could also pose potential harm.

Unfortunately, amongst the popular biomedical Natural Language Processing (NLP) datasets for evaluating LLMs, none of them have been specifically prepared for resourceconstrained settings as found in Low- and Low-Middle-Income countries (LMICs) Zhou et al. (2023). Thus, though a few models achieve high scores when evaluated on these datasets, their translational value in everyday clinical scenarios in resource-constrained cannot be readily ascertained.

Moreover, in resource-constrained settings, the ability to obtain, create, or utilize specialized, domain-specific models is greatly limited by factors such as cost. Previous research has demonstrated that employing advanced prompting strategies with generalized LLMs can yield outcomes surpassing those of specialized medical LLMs, as evidenced by MedPrompt. Nori et al. (2023). If generalized models, which are often more accessible to broader populations, can be optimized to achieve comparable or superior performance to specialized medical LLMs through simpler prompt engineering techniques, then developers in resource-constrained settings can leverage this opportunity to create effective yet cost-efficient applications tailored to their environments.

In this study, our objective is to contribute to the scant knowledge base regarding LLM applications for clinical scenarios in low- and middle-income countries (LMICs), while also serving as a reference for future, more comprehensive research. Specifically, we aim to assess the suitability of certain selected generalized LLMs as clinical decision support tools for managing medical emergencies in LMICs. This study forms part of a broader research and development initiative aimed at eventually implementing such tools in resource-constrained settings.

### Generalizable Insights about Machine Learning in the Context of Healthcare

1. To the best of our knowledge, this is the first comparative study with clinician evaluation to investigate the effectiveness of state-of-the-art generalized LLMs in providing first aid instructions for resource-limited settings with diverse prompting strategies.
2. We test and compare two uncommon prompting strategies, namely self-questioning prompting: which elicits informative questions pertinent to the clinical scenarios at hand to improve the model’s respone, and a stacking of self-questioning prompting and the more popular chain-of-thought prompting technique(SQCT).
3. We demonstrate that prompting techniques are not one-size-fits-all, i.e. different prompting techniques might elicit better responses in different models.
4. Our findings emphasize the need for continuous human expert verification in evaluating the clinical utility of LLM-based health applications.

## 2. Related Work

In this section, we review the relevant literature on large language models applied to clinical language understanding tasks in healthcare, as well as existing prompting strategies.

### 2.1 LLMs in Healthcare for Resource-Constrained Regions

Prior studies have shown that though there are vital concerns to be addressed, the general consensus is that LLMs hold immense potential in improving healthcare delivery when they are incorporated in various capacities such as: in automation of administrative tasks, clinical decision support tools, virtual health assistants, screening tools, health trackers, clinical language translation tools, medical research and health education tools Goodman et al. (2023) Tripathi et al. (2024) Sallam (2023) Abu-Jeyyab et al. (2023). These functions can augment the limited financial, logistical and human resources available in LMICs Gangavarapu (2023). Initial studies on clinician perception on the usefulness of a combination of OpenAI’s “gpt-3.5-turbo / “gpt-4” and Retrieval Augmented Generation (RAG), as a health education tool in India, an LMIC, revealed that though clinicians believed the tool held potential, they were generally not satisfied with its performance Al Ghadban et al. (2023). In that study, the authors identified the need to enhance the contextual and cultural relevance of the models responses. Another comparative study of a clinician evaluation of Almanac, an LLM framework based on OpenAI’S “text-davinci-003” combined with RAG, versus ChatGPT reveals that though clinicians rated Almanac’s answers as safer and more factual, they still preferred ChatGPT’s answers Zakka et al. (2024). However, this study does not reveal whether the clinicians shared their perspective on the usefulness of any of the models for everyday clinical scenarios, neither does it capture the perspectives of clinicians who practice in LMICs.

### 2.2 Prompting strategies

#### Few-shot prompting

Few-shot prompting involves giving the model demonstrations of the task at hand usually in the format of “User:” followed by an example user input and “Model/Assistant:” followed by an expected model response. When only one demonstration is given, it is termed one-shot prompting Brown et al. (2020). When five demonstrations are given, it is termed five shot prompting (5SP), and when no demonstrations are given it is termed zero-shot prompting.

A study assessing the performance of an LLM on health-related tasks found that fewshot prompting led to greater increases in the model’s accuracy and decreases in error as compared to zero-shot prompting, and even few-shot supervised training Liu et al. (2023a). A study comparing the performance of a combination of various generalized LLMs and prompting techniques on questions in the thoracic surgery domain revealed that few-shot prompting outperformed zero-shot prompting. Furthermore, five-shot prompting consistently outperformed one-shot prompting Li et al. (2023). Another study comparing zeroshot prompting to five-shot prompting across various clinical and biomedical tasks, also found that overall, five-shot prompting exceeded zero-shot prompting Labrak et al. (2023).

#### Chain-of-thought prompting

Chain-of-thought prompting (COT) involves leading the model to break down complex tasks into intermediate steps, so it can follow a step-bystep approach to arrive at its response. This approach has been touted to improve model reasoning and performance Wei et al. (2022). A study comparing COT with the few-shot prompting strategy using the Flan-PaLM 540B model across a range of benchmark medical question-answering tasks, did not find substantial improvement over few-shot prompting Singhal et al. (2023).

#### Self-questioning

Self-questioning prompting (SQP), is a relatively new strategy which elicits improved questions based on the original user input to generate additional context for the original input, with the aim of generating a better model response. This approach has been shown to elicit better model performance compared to 5SP across a few clinical language understanding tasks Wang et al. (2023). However, this approach has not been tested in multiple end-user scenarios.

#### 2.3 Model selection and parameters

A comparative study on clinical text summarization across eight LLMs and various model temperatures revealed that GPT-4 and the lowest temperature (0.1) yielded the best results Van Veen et al. (2024). In another comparative LLM study on various clinical benchmark datasets spanning multiple tasks, GPT-4 outperformed GPT-3.5 and Bard [K]. Another study comparing the performance of Claude-instant-v1.0, GPT-3.5-Turbo, Commandxlarge-nightly, and Bloomz, in three clinical specialties found that different models performed better on different metrics Wilhelm et al. (2023). Another study comparing the performance of GPT-3.5, PaLM-2, Claude-2, and LLaMA-2 on 6 biomedical tasks found that the best performing LLM varied across various tasks Jahan et al. (2024).

### 2.4 Methods For Evaluating the Performance of LLMs in clinical tasks

A study evaluating the performance of various LLMs on answering clinical questions in Radiology measured their performance against two datasets in Radiology and using Recall@1, Recall@2 and Recall@L as metrics Liu et al. (2023b). This can be a useful evaluation rubric but may not be applicable to clinical domains/scenarios in which different contexts might necessitate different approaches to diagnosis and management. Another publication proposes “Artificial-intelligence Structured Clinical Examinations” as an evaluation framework that uses agent-based simulations to mimic real world clinical scenarios Mehandru et al. (2023). Though promising, this framework has not been thoroughly studied or implemented. In another study evaluating a radiological vision-language model’s output, the authors employed both automated evaluation via the popular NLP metrics such as BLEU score and Rouge-L, and human expert evaluation, noting that the former could not properly assess for factual correctness and consistency – properties that are vital for clinical utility Tanno et al. (2024). Another study on LLM outputs for medical evidence summarization tasks also employed both automatic and human evaluation. They defined summary quality based on coherence, factual consistency, comprehensiveness, and harmfulness Tang et al. (2023). The researchers concluded that automatic metrics often do not strongly correlate with the quality of summaries. Another study on the performance on medical LLMs on clinical application tasks also emphasized the limitations of automated metrics to measure clinical utility and employed human experts to rate model answers on: agreement with scientific and clinical consensus, the presence of incorrect content, the omission of content, the extent of possible harm, the likelihood of harm, and possible bias in answers Singhal et al. (2023). A proposed evaluation framework for the implementation of Artificial Intelligence systems into healthcare settings proposes that AI applications be assessed on three main components: adoption, capability, and utility, with 15 subcomponents such as safety and quality, non-maleficence, generalizability and contextualization Reddy et al. (2021). In a recent study, researchers assessed the output of a medical LLM by asking human experts to assess the model’s responses on a 17-metric evaluation rubric along five axes: accuracy, safety, fairness, interpretation and communication Bosselut et al. (2024). Another study on clinical text summarization compared LLM summaries to summaries by medical experts on correctness, conciseness and completeness and found that NLP metrics correlated poorly with clinician preferences, emphasizing that NLP metrics are inadequate in assessing clinical readiness of LLMs Van Veen et al. (2024).

## 3. Methods

In this section, we detail our experimental setup and analytical techniques.

### 3.1 Selection of medical scenarios and clinician evaluators

We simulated 10 common emergency medical scenarios in Ghana collaboration with a panel of 12 clinicians led by an emergency medicine specialist with 12+ years of practice experience in Ghana. The clinical scenarios featured a diverse range of demographic characteristics with patient ages spanning from 5 months to 50 years and featured an equal distribution of male and female patients. The clinical scenarios also cut across major clinical specialties including Internal Medicine, Obstetrics and Gynaecology, General Surgery and Paediatrics. These scenarios were then incorporated into prompt templates that were fed into models and evaluated by a different set of 22 clinicians. Clinician evaluators were selected via a local clinician network, from diverse practice locations within Ghana and based on their familiarity with the locations, contexts, and clinical scenarios. All clinician evaluators had at least 2 years of clinical practice experience in resource-constrained settings in Ghana, specifically as the first point-of-call in the hospital in managing medical emergencies in Ghana. It is expected that they possess sufficient knowledge and skills to deliver, at a minimum, first aid in the selected medical scenarios.

### 3.2 Model selection

We tested Open AI’s GPT-4-0125 Preview, via the OpenAI Assistants Platfom, Gemini 1.5 Pro via Google AI Studio, and both Claude Sonnet and Opus via the Anthropic Console. These models were selected based on performance on popular benchmarks, their ranking on the LMSYS Chatbot Arena Leaderboard as of 13th April 2024, availability of API, ease of access and for further cost-benefit ratio comparative analysis Zhou et al. (2023) Chiang et al. (2024). We did not select similarly-ranked open-source medical LLMs because of the computational resources required to run/access them, for example, advanced GPUs.

### 3.3 Parameter Tuning

The GPT-4-0125 Preview model was set to the default temperature of 1 due to the difficulty in tweaking the temperature at the time of running the tests (reference documentation was only recently updated to make this option readily accessible). Similarly, the Gemini 1.5 Pro model was tested at its default temperature of 2 as this could not be easily modified. The Claude Sonnet and Claude Opus models were both tested at a temperature of 0. This approach was to generate deterministic responses as often as possible due to the critical nature of the use case and was guided by findings from internal tests and findings from research studies detailed in Section 2.3 above.

### 3.4 Prompt Engineering

Four prompt engineering techniques were tested: In-Context Learning with 5-shot prompting (5SP), chain-of-thought prompting (Co**T**), self-questioning prompting (SQP), and a stacking of self-questioning prompting and chain-of-thought (SQC**T**). A general prompt structure was provided, and the prompt modified to suit the technique being tested. The general prompt structure consisted of the system message/instruction and the clinical scenario. The last part of the prompt consisted of a modification based on the prompt technique being tested. Figure 1 shows the general prompt structure, and an example prompt across the four techniques tested. The prompt modifications are highlighted.

**Figure 1:**
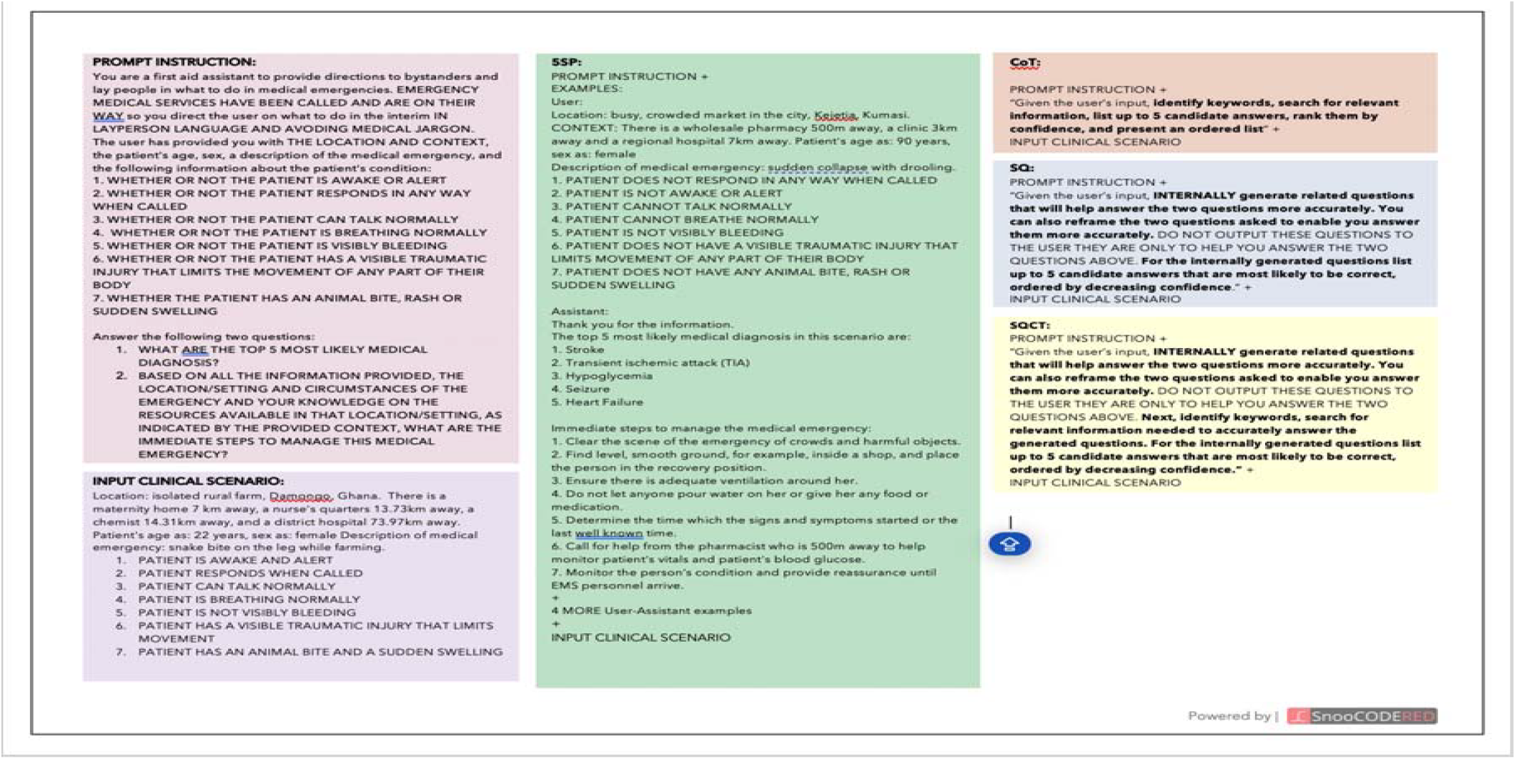
Differences in prompt based on prompt technique.

### 3.5 Distribution of Prompt-Response Pairs to Evaluators

Each of the four models tested was paired with each of the four prompt techniques yielding 16 model-prompt combinations. Eight model-prompt combinations were run twice, and eight model-prompt combinations were run thrice, this produced 40 test runs. For each of the 10 clinical scenarios, four model-prompt combinations were tested, and their responses presented for evaluation. Each clinician was assigned to blindly evaluate 20 prompt-response pairs with a total of 440 complete evaluations expected. Figure 2 shows a visualization of the exact model-prompt technique pairs and the number of times they were run.

**Figure 2:**
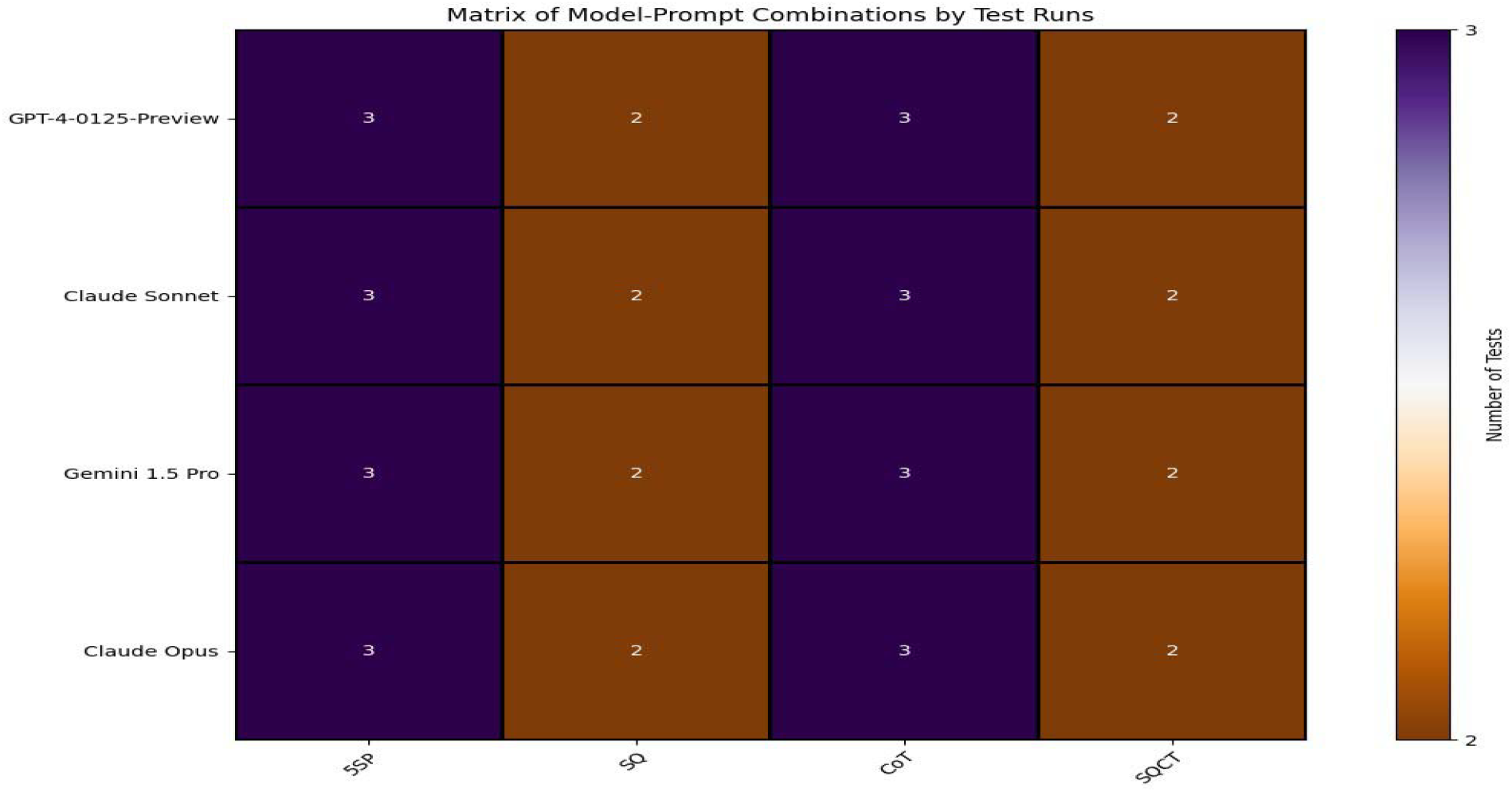
Model-Prompt Technique Combinations Per Number of Runs.

### 3.6 Response Evaluation

Clinicians were told they were ranking AI models but were not told which models were being ranked and which response was generated by which model. Each prompt was followed by 4 responses and each response was graded on accuracy, conciseness, helpfulness and then an overall score. Grading was done on a 10-point Likert scale, with 0 representing “Totally Unsatisfactory” and “Totally Satisfactory”. At the end of the four responses to each prompt, a comment box was provided, and evaluators asked to input any additional comments they had about the prompt and responses. An example evaluation form is shown in the Appendix A.

### 3.7 Collection and Analysis of Evaluation Reports

Evaluation reports were collected via an online form. Data preparation, quantitative analysis and associated data visualizations were performed in Microsoft Excel Version 16.83 and with Python 3.11 Pyt. The Real Statistics Resource Pack 14 was used for Interrater Reliability Analysis rea. For qualitative analysis, evaluators’ comments were compiled as text in a document and coding was performed using Taguette 1.4.1-40-gfea859715 Rampin and Rampin (2021). Subsequently, thematic analysis and data visualization were performed in Python 3.11. For the data wrangling process 12 entries had no ratings across the evaluation rubric and were taken out leaving a total of 428 entries for the subsequent analysis. Furthermore for the correlation analysis, 2 ratings with no overall scores were taken out. Mean imputation was then performed for the following missing values: 3 missing ratings under “Conciseness”, 1 under “Accuracy” 2 under “Safety” and 1 under “Helpfulness”.

## 4. Results

In this section, we present a comprehensive quantitative analysis of the performance of the LLMs, the prompts and the LLM-prompt pairs based on clinician evaluation. We began by comparing the performance of the models, in terms of accuracy, conciseness, helpfulness and the overall score of their responses. Next, we compared the various prompt techniques also on accuracy, conciseness, helpfulness and the overall score. **T**hen we compared the modelprompt technique pairs based on the same rubric. Next, we investigated the correlation between accuracy, conciseness and helpfulness of responses with the overall score. Next, we assessed interrater reliability. Finally, we provided a qualitative analysis of clinician comments.

### 4.1 Comparison of Model Performance Based on Clinician Rating

The four LLMs: GPT-4-0125-Preview, Gemini 1.5 Pro, Claude Sonnet and Claude Opus were compared based on response conciseness, accuracy, safety, helpfulness and overall score as graded by evaluators. The mean ratings was calculated for each model across the four different prompt techniques tested. All the LLMs had the lowest scores in response conciseness and the highest scores in response helpfulness as compared to the other categories. Claude Sonnet scored the highest scores across the evaluation rubric. Table 1 shows the mean rating scores with the highest in each category highlighted in bold.

**Table 1:** Mean Clinician Rating of Selected LLMs on a 10-point Likert scale.

| Short Title |  |  |  |  |  |
| --- | --- | --- | --- | --- | --- |
| Model | Conciseness | Accuracy | Safety | Helpfulness | OverallScore(s.d.) |
| GPT-4-0125-PREVIEW via OpenAI Assistant | 5.23 | 6.79 | 7.09 | 7.26 | 6.94 (1.58) |
| GEMINI 1.5 PRO | 7.11 | 6.98 | 7.06 | 7.18 | 7.15 (1.58) |
| **CLAUDE SONNET** | **7.48** | **7.49** | **7.7** | **7.74** | **7.7 (1.3)** |
| CLAUDE OPUS | 6.35 | 7 | 7.15 | 7.22 | 7.28 (1.76) |

**Table 2:** Comparison of Prompt Techniques Per Response Rating.

| Prompt Technique | Conciseness | Accuracy | Safety | Helpfulness | Overall Score (s.d.) |
| --- | --- | --- | --- | --- | --- |
| 5SP | 6.62 | **7.4** | **7.6** | **7.67** | **7.47 (1.43)** |
| CoT | 6.32 | 6.74 | 6.83 | 6.83 | 7.06 (1.74) |
| SQP | **6.88** | 7.1 | 7.36 | 7.36 | 7.25 (1.4) |
| SQCT | 6.36 | 7.05 | 7.1 | 7.1 | 7.23 (1.76) |

### 4.2 Comparison of Prompt Techniques Based on Mean Clinician Rating

The four prompt techniques tested were also compared based on the rating of the model responses generated by the prompts. The mean ratings for each prompt technique were calculated across all the four models tested. Again, the lowest scores were for response conciseness and the highest for response helpfulness. Response safety was almost at par with response helpfulness. **T**able 2 shows the mean rating scores with the highest in each category highlighted in bold.

### 4.3 Comparison of Model-Prompt Technique Combinations based on MeanClinician Rating

The 16 model-prompt technique combinations were compared across the evaluation rubric and the findings summarized in Table 3 and Table 4. Claude Sonnet + SQCT produced the best rated responses generally, whereas Gemini 1.5 Pro + Co**T** or GP**T**-4-0125 Preview + Co**T** produced the least-rated responses across the evaluation rubric.

**Table 3:** Best-Performing Model-Prompt Technique Cobination Per Evaluation Metric.

| Evaluation Rubric | Highest-Ranked Model-Prompt Technique | Mean Score (s.d.) |
| --- | --- | --- |
| Conciseness | Claude Sonnet + SQP | 8.24 (1.51) |
| Accuracy | Claude Sonnet + SQCT | 7.95 (1.29) |
| Safety | Claude Sonnet + SQCT | 7.9 (1.34) |
| Helpfulness | Claude Sonnet + SQCT | 8.14 (1.28) |
| Overall Score | Claude Sonnet + SQCT | 8.05 (1.17) |

**Table 4:** Worst-Performing Model-Prompt Technique Cobination Per Evaluation Metric.

| Evaluation Rubric | Lowest-Ranked Model-Prompt Technique | Mean Score (s.d.) |
| --- | --- | --- |
| Conciseness | GPT-4-0125-Preview + CoT | 4.63 (2.52) |
| Accuracy | GPT-4-0125-Preview + CoT | 6.28 (2.29) |
| Safety | Gemini 1.5 Pro + CoT | 6.43 (1.7) |
| Helpfulness | Gemini 1.5 Pro + CoT | 6.61 (1.85) |
| Overall Score | Gemini 1.5 Pro + CoT | 6.5 (1.62) |

### 4.4 Correlation between Response Conciseness, Accuracy, Safety andHelpfulness with Overall Score

A linear regression model was fitted to determine the relation between response conciseness, accuracy, safety and helpfulness to the overall score to determine which of these factors was closely related with the overall score. As shown in Figure 3 the helpfulness of the response had the highest correlation with the overall score. The response safety had the second highest correlation. Appendix B shows the suitability of the linear regression model for the correlation analysis.

**Figure 3:**
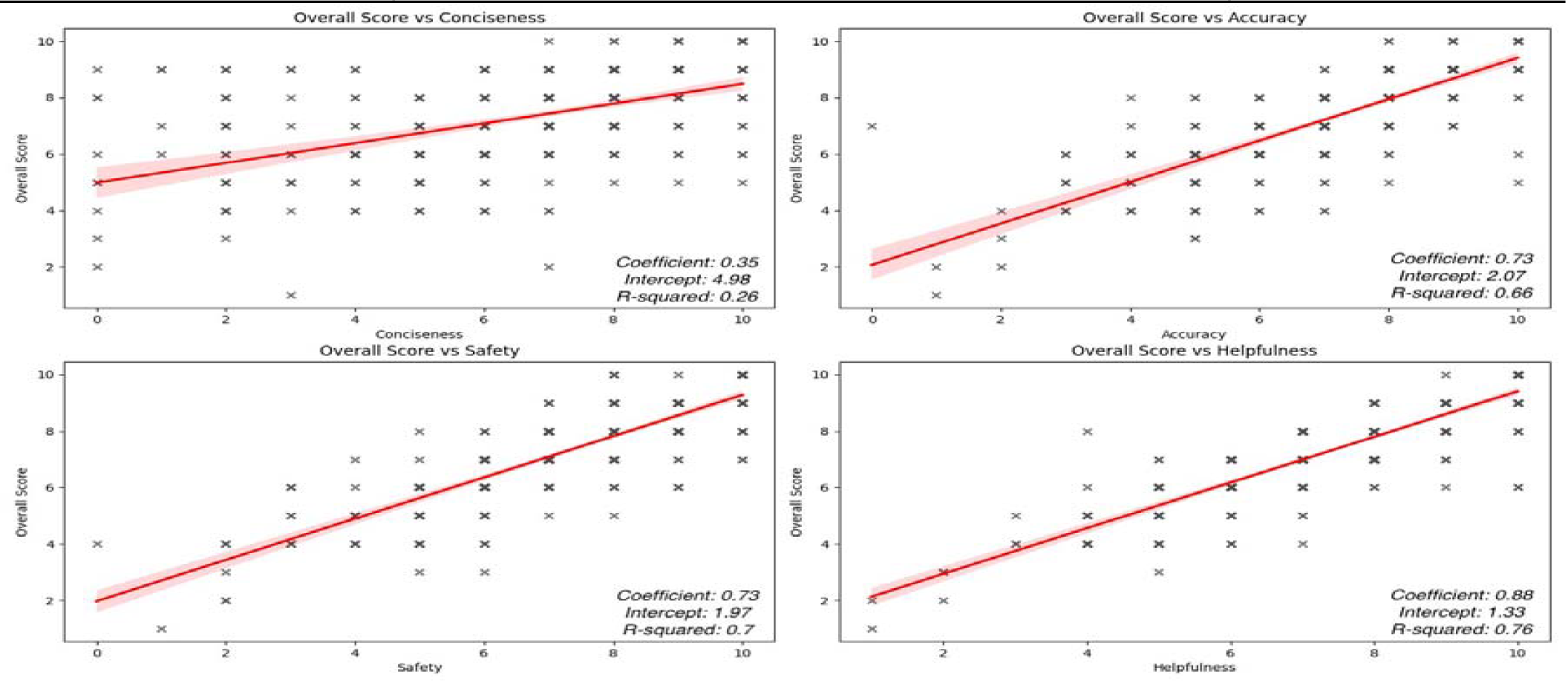
Regression analysis of Overall Score vs Conciseness, Accuracy, Safety and Helpfulness.

### 4.5 Interrater Reliability

To measure the agreement between evaluators on the overall score, we use Gwet’s AC2 coefficient Gwet (2014). Gwet’s AC2 score across all the ratings for overall score was 0.79 corresponding to a moderate to strong agreement between raters Gwet (2014). Appendix C shows the full results of the Interrater Reliability Analysis.

### 4.6 Qualitative Analysis

Majority of clinician comments affirmed the clinical accuracy of the responses. The second most common theme in the comments was about clinicians adding first aid steps they perceived as missing from the model’s response. Clinicians also shared a few comments about diagnosis they were expecting to see that were missing from the responses. Few comments expressed displeasure or dissatisfaction in the responses affirming the relatively high rating scores given by clinicians. Table 5 describes the codes used to encapsulate the themes of clinician comments and Figure 4 shows the frequency of codes in all of the comments left by clinicians. Figure 5 is a word cloud that gives an overview of the most common words and phrases in the additional first aid instructions provided by clinicians to make the responses better. The more frequently occurring a word or phrase is, the more prominent it is in the word cloud.

**Table 5:** Coding of Evaluators’ Comments.

| Code | Description | Example comment |
| --- | --- | --- |
| EmphasisOnMedicalExpertise | Evaluator emphasizes a need to seek further medical management by an expert. | "Medical evaluation is recommended" |
| AccurateResponse | Evaluator commends the clinical accuracy of the response | "All responders were quite accurate with their differentials and their ways of delivering first aid." |
| InaccurateResponse | Evaluator comments that the response is not clinically accurate | "Responder B2 however deviated" |
| QuickTransport | Evaluator emphasizes the need of quick transportation | "arrange a quick transportation" |
| AdditionalFirstAidStep | Evaluator added at least one additional first aid step. | "Some other measures will include shouting for help from onlookers" |
| Misdiagnosis | Evaluator disagrees with at least one of the top 5 medical diagnosis given. | "meningitis and diabetic ketoacidosis in my opinion should not be among the topmost diagnoses." |
| HospitalPrep | The evaluator emphasizes informing the hospital before patient is transported. | "Immediate communication with the hospital is necessary to ensure adequate preparation" |
| SatisfactoryResponse | Evaluator is pleased with response. | "Good responses so far." |
| Setting-Deaf Response | The response is not appropriate for the setting | "it is quite unrealistic for a layman to have oxygen and AED at home as stated Responder C" |
| MissingDiagnosis | Evaluator emphasizes that some expected diagnosis was missing in response. | "In a country like Ghana and especially in rural settings, malaria should be a differential diagnosis for this scenario" |

**Table 6:** Interrater Reliability Analysis Results.

Gwet's AC2 Analysis
|  |  |
| --- | --- |
| Coefficient | 0.78710729 |
| Standard error (subject) | 0.02555862 |
| Confidence interval (lower border) | 0.73263039 |
| Confidence interval (upper border) | 0.8415842 |
| Standard error (total) | 0.05477784 |
| Confidence interval (lower border) | 0.67035109 |
| Confidence interval (upper border) | 0.9038635 |

**Figure 4:**
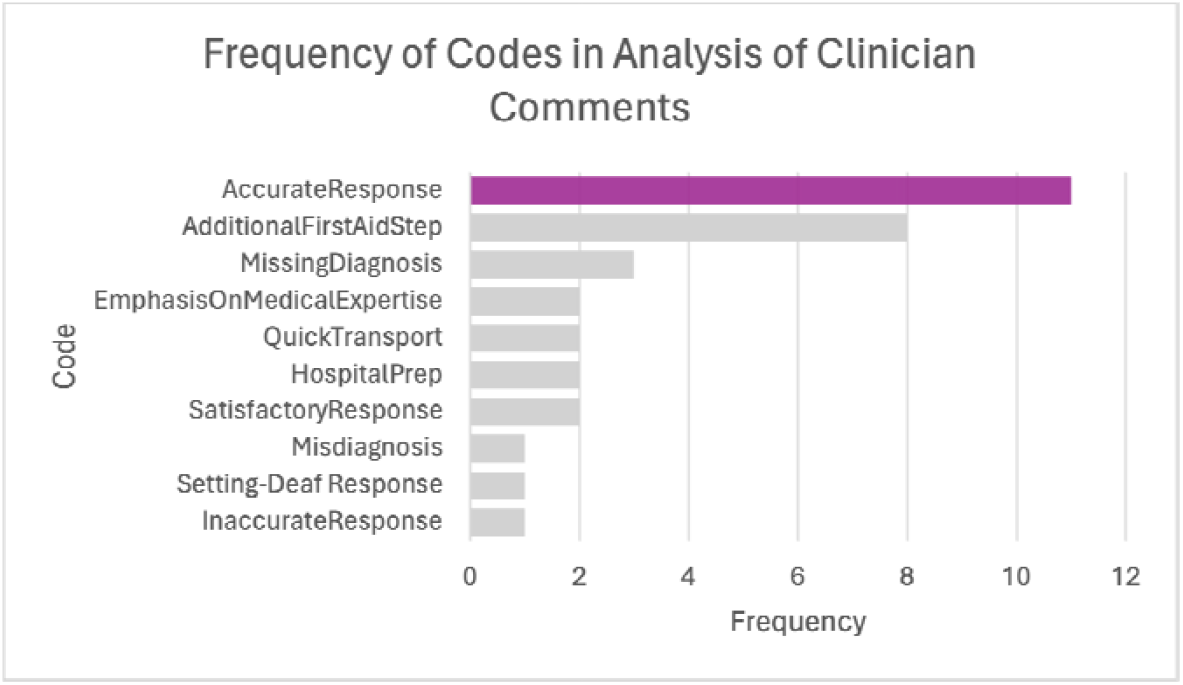
Frequency of Codes in Evaluators’ comments with the most frequently occurring code highlighted.

**Figure 5:**
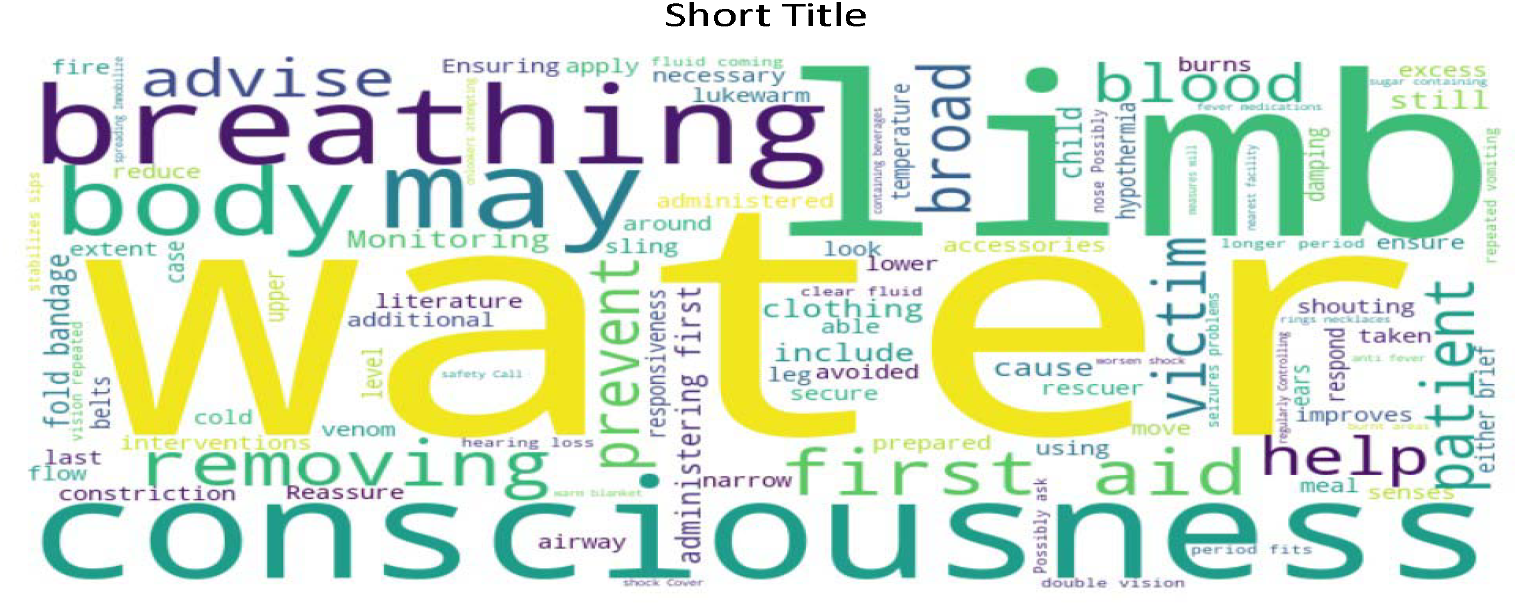
Visualization of Evaluators’ comments under the AdditionalFirstAidStep code.

#### Clinician Opinion on LLMs in First Aid Decicion Support Tools

Clinicians were asked to share their opinion on LLMs being used in first aid decision support tools by answering Yes/No to the following question, and to leave any additional comments in a textbox provided beneath it: *A first aid tool is being developed for call centres. Bystanders/laypersons can call the centre and be directed by a responder on what to do in the interim as they wait for EMS. Do you think at least one of the models you have assessed today will be useful if incorporated into this tool?* All 22 clinicians responded Yes without any additional comments.

## 5. Discussion

The results from both quantitative and qualitative analysis show that clinician evaluators were generally satisfied with the diagnosis and first aid instructions outputted by the best performing generalized LLMs. This performance by the LLMs is notable considering that they were neither specially built medical LLMs nor had they had any prior pretraining or finetuning for the tasks. Also, the prompting strategies tested were relatively simple as compared to more sophisticated state-of-the-art techniques like MedPrompt Nori et al. (2023). The best performing model-prompt technique combination in our study, achieved a mean ranking score of 8.05/10 which is promising. This is a promising finding for developers of LLM-based health applications in resource-constrained settings where the ability to create more specialized, domain-specific models and/or to run them is greatly limited. Applications of this nature have the potential to reduce glaring disparities in healthcare delivery in countries like Ghana. For example in Ghana, per estimates by the World Health Organization (WHO), for every ten cases referred by the national ambulance service in Ghana in 2022, one did not receive emergency support either due to lack of ambulances or bed space in receiving hospitals for Africa (2023). For those who receive care at the hospitals, the reported doctor to patient ratio is 1:6500, way below the WHO’s recommendation of 1:1000 for Africa (2022) Mullan and Bryant (1984). To exacerbate the problem, 81.3% of doctors are in only 5 regions, and over 60% of doctors are in only 5 teaching hospitals in the country for Africa (2022). If LLM-based applications can be developed to offer dependable first aid guidance tailored to the local context, then in regions of Ghana with limited ambulances and few clinicians, assistance can be extended to emergency victims by laypersons and minimally trained first aid providers. This assistance can be provided while efforts are made to access the services of more qualified personnel.

An example of such an application is the SnooCODE Red app being developed in Ghana. Figure 6 shows a version of the app in development.

**Figure 6:**
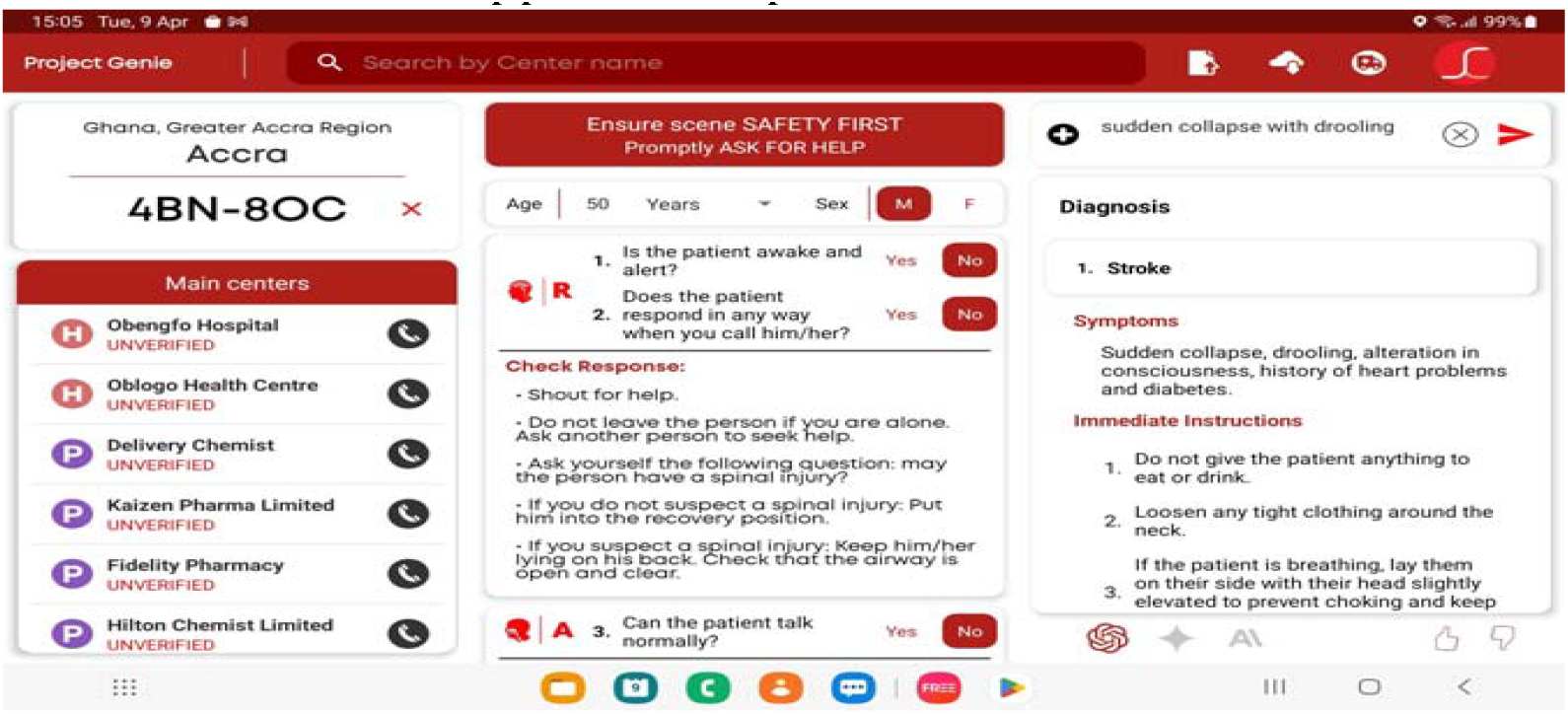
A Screenshot of the SnooCODE RED app under development.

**Figure 7:**
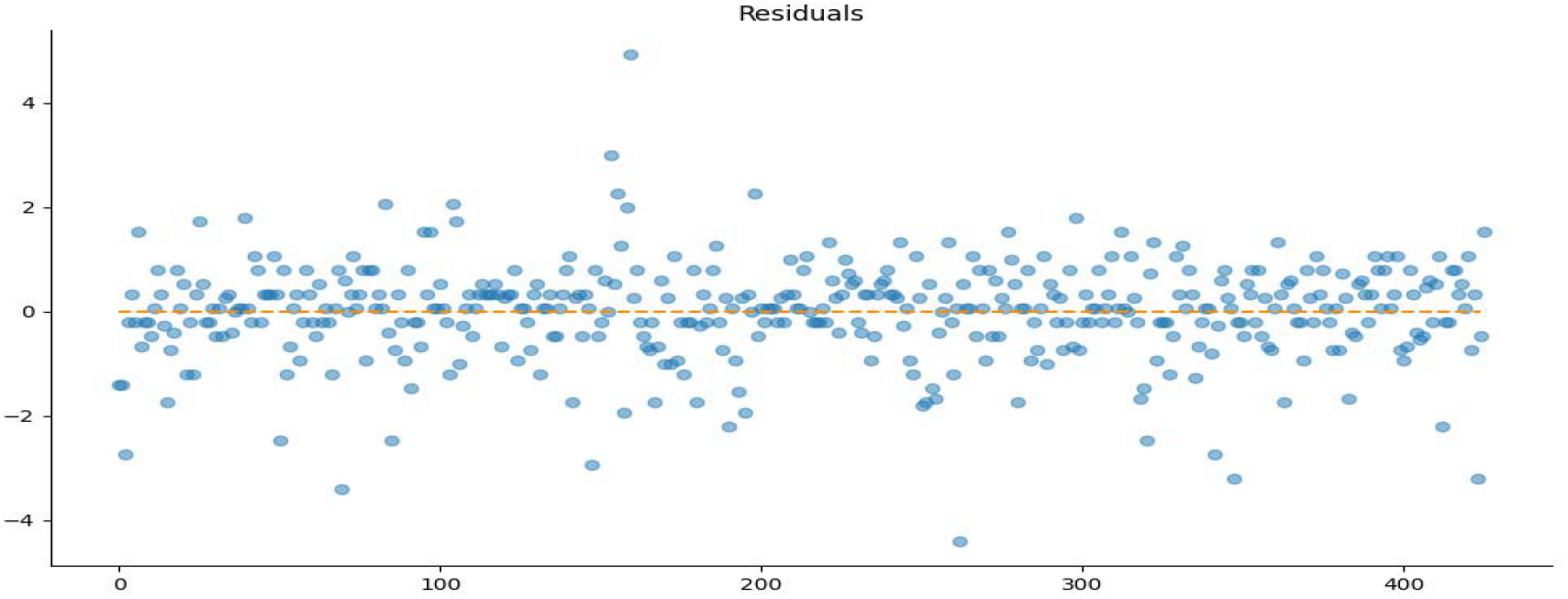
Test for Homoscedasticity: Linear Regression of Overall Score vs Accuracy.

**Figure 8:**
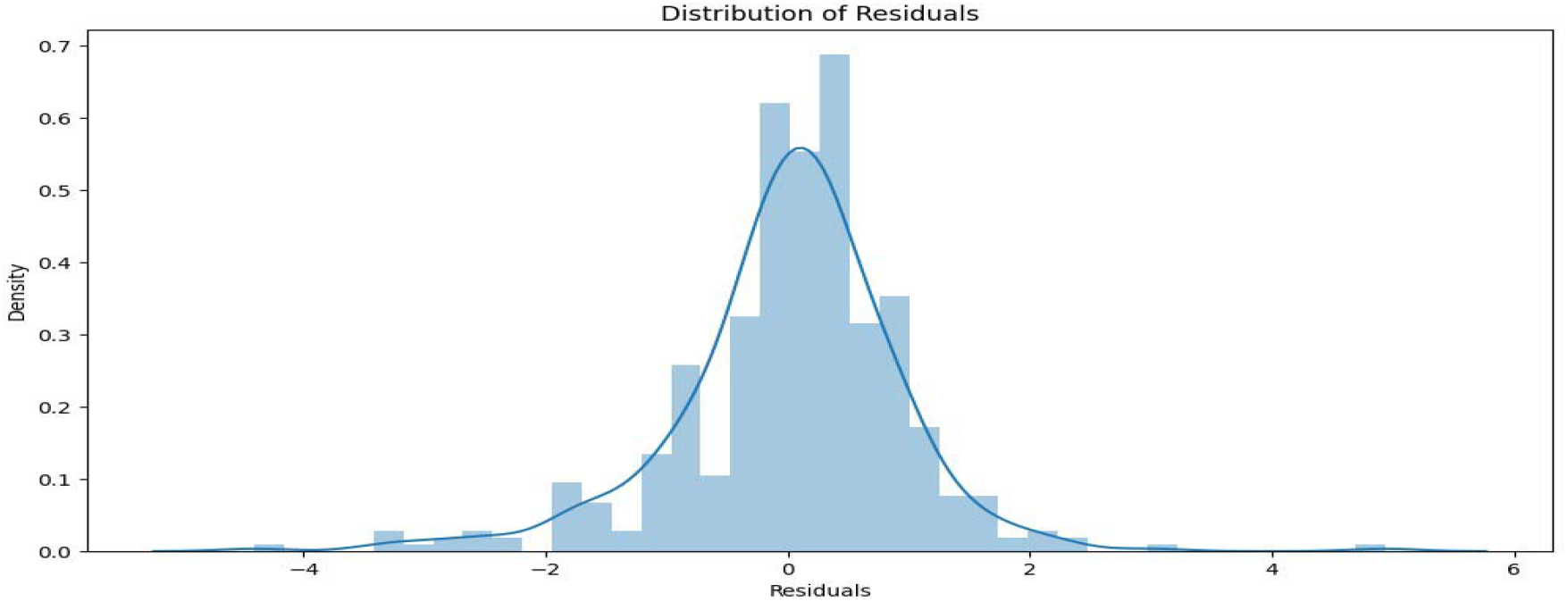
Distribution of Residuals: Linear Regression of Overall Score vs Accuracy.

**Figure 9:**
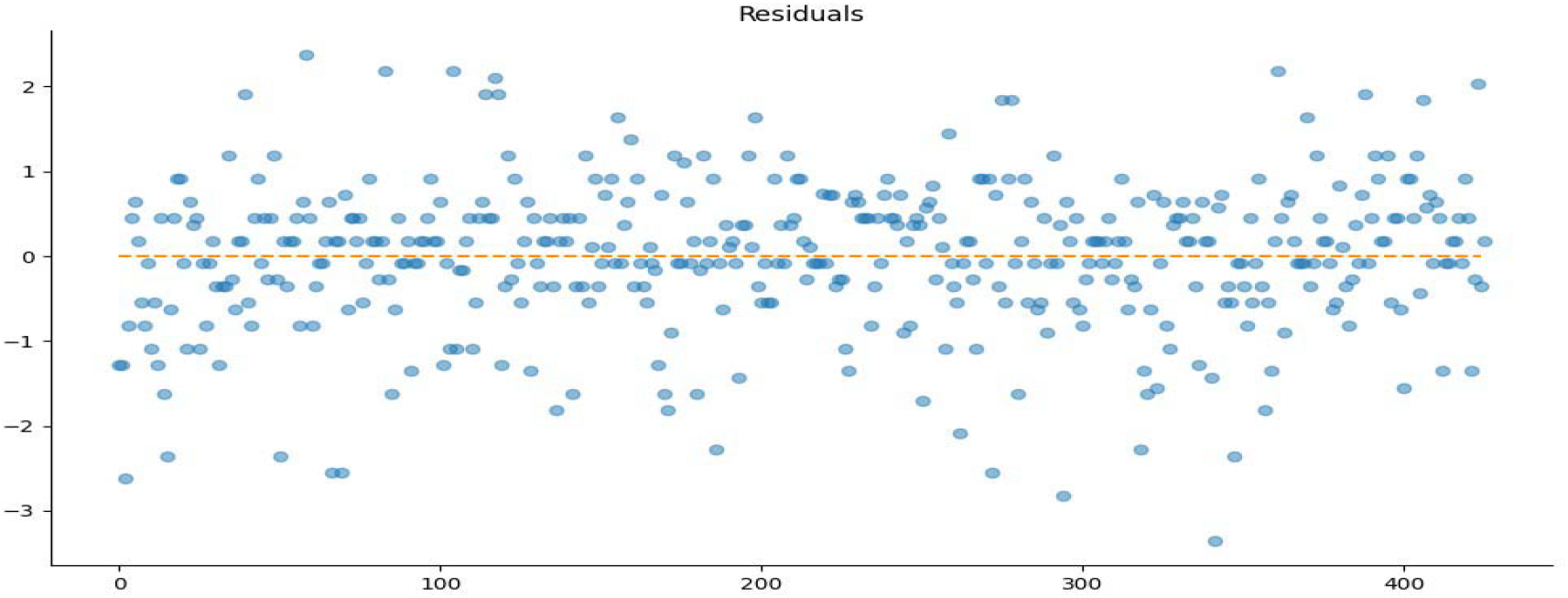
Test for Homoscedasticity: Linear Regression of Overall Score vs Safety.

**Figure 10:**
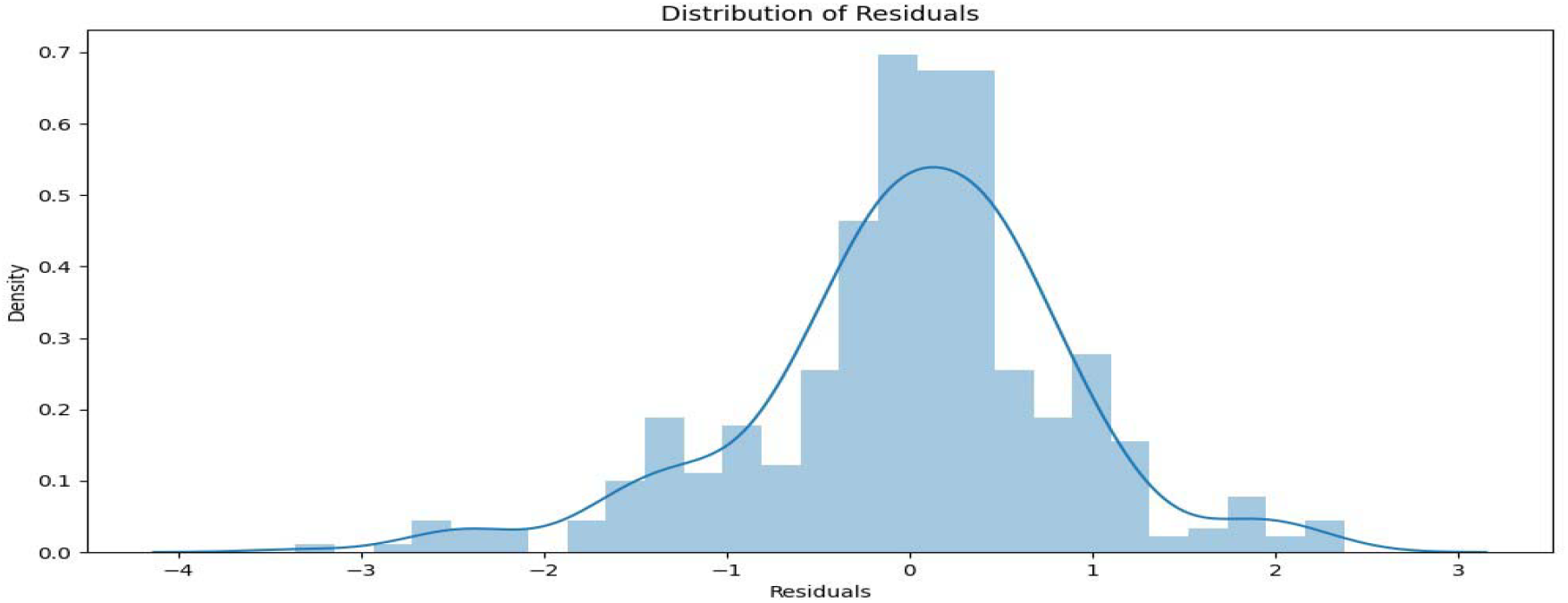
Distribution of Residuals: Linear Regression of Overall Score vs Safety.

**Figure 11:**
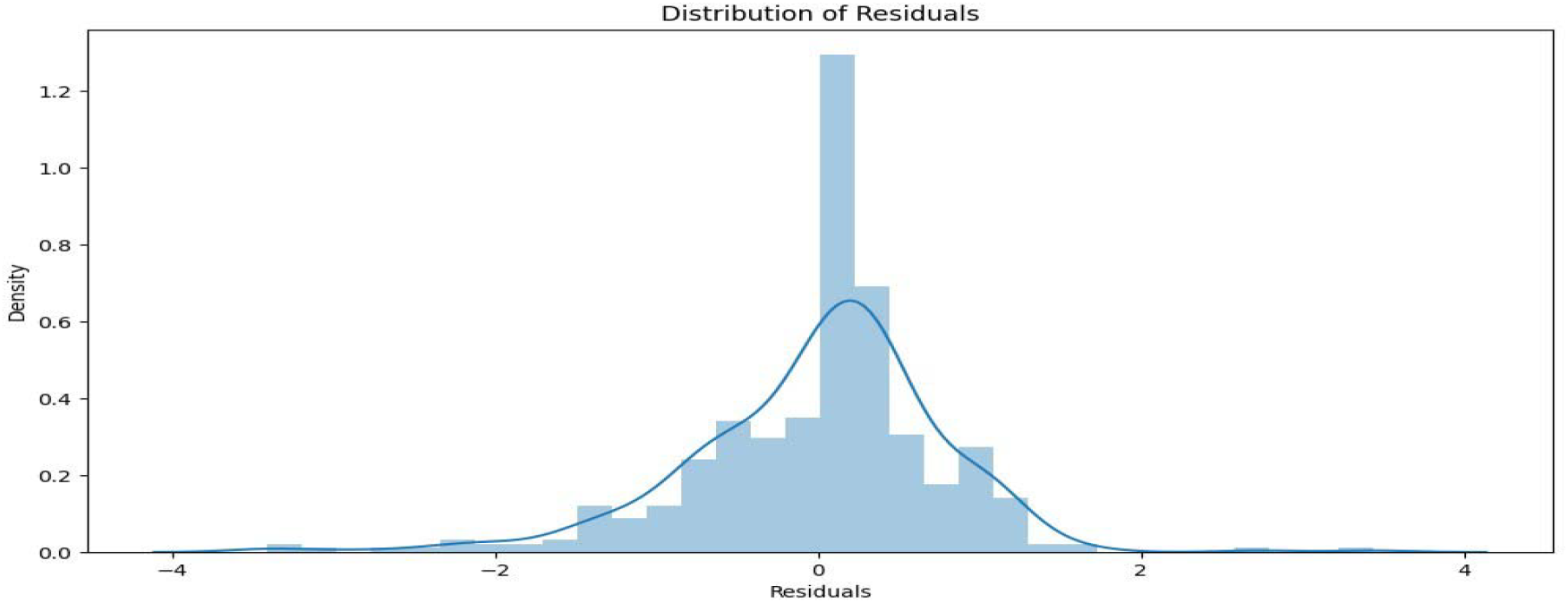
Distribution of Residuals: Linear Regression of Overall Score vs Helpfulness.

Another interesting finding is that more expensive LLMs may not necessarily have better performance than less expensive models, at least for clinical tasks. In the same vein, higher-ranked models on popular leaderboards may not necessarily perform better for clinical use. Though leaderboards and model scores on conventional benchmarks may guide developers in choosing models, for developers of LLM-based health applications, it is important to continuously and thoroughly test models for the intended use case and not rely on mainstream information only.

It was also interesting to see the results from the testing of the Self-Questioning Prompt technique. Its authors noted that it elicited better model responses compared to the more popular Chain-of-Thought prompting technique and our findings seemed to confirm this claim Wang et al. (2023). We also suggest combining Self-Questioning and Chain-ofThought prompting techniques and testing both approaches to see which performs better.

It was interesting to see that some prompting techniques elicit better responses when paired with some models but not when paired with others. Even though five-shot prompting seemed to be the best prompting technique overall, for Claude Sonnet, the stacked Self-Questioning and Chain-of-Thought prompting technique seemed to yield better responses for the model. Further experiments need to be performed to see if this behaviour is consistent and why that might be the case. If this behaviour proves consistent, it might be welcoming news in cases where as five-shot prompting may use more tokens than the other prompting techniques.

The results from the qualitative analysis give the impression that clinicians were in favour of the LLMs being used as first aid decision support tools. It was impressive that the evaluators unanimously affirmed the usefulness of LLMS as first aid decision support tools. However, as noted by the frequent additional first aid steps given by clinicians, thorough work has to be done before such a tool is tested in real-world settings. It was interesting to analyze the common phrases in the additional first aid steps given by clinicians. The most commonly occurring word “water” is not surprising given how clean water is vital for sustaining life. Access to clean, potable water can be a challenge in the resource-constrained settings simulated in this experiment. Other common phrases like “consciousness”, “breathing” and “limb” appear in almost the exact order in which they are to be assessed in the Advanced Trauma Life Support protocol used by emergency responders and physicians. Their prominence in clinician comments show the importance of assessing and correcting defects in these areas during first aid provision. It is clear that an approach of continuous iteration and collaboration with users and medical experts must be adopted in building LLM-based applications for healthcare. As more AI researchers devise methods to automatically assess model performance, caution must be exercised, particularly for critical uses like this one. For applications in LMICs especially, conventional LLMS may fall short in capturing the nuances of resource-constrained settings and thus it is important to involve stakeholders from such settings in the development process to ensure that LLM-based health applications for such settings can have real translational value.

### Next Steps

Next steps include improving model responses based on the findings from the study and from clinician feedback. Also, a similar study is being conducted involving more experienced clinician evaluators, and subsequently a larger cohort of clinicians. We hope that the findings from this study can provide some direction for more extensive research that would help alleviate the research gap on LLM applications in LMICs. ltimately, our goal is to develop LLM-based solutions that hold clinical value for low- and middle-income countries (LMICs).

### Limitations

Though our study provides some insight on clinician perception of LLM usefulness in first aid decision support tools for resource-constrained settings, there are obvious limitations with this study. Firstly, a larger cohort of clinical scenarios would have provided a better assessment of the performance of the various models and prompt techniques. Also, specially trained medical LLMs might have provided better responses as compared to generalized LLMs for our use case. Again, it would be helpful to have gotten annotations from more experienced clinicians, e.g. from emergency medicine specialists with many years of practice in Ghana. Again, the experiment could have been conducted for various resource-constrained areas outside of Ghana to enhance the generalizability of the findings. Furthermore, ethical and legal considerations of using the LLMs for provision of first aid advice are not addressed in this study. Future work should explore these aspects to expedite the application of LLMs in resource-constrained settings.

## Data Availability

All data produced in the present study are available upon reasonable request to the authors

## Appendix A.

A sample form for collection of clinician responses can be found at this link: . It also hows a section of the clinical scenarios simulated.

## Appendix B.

The figures below demonstrate the suitability tests performed for the linear regression.

**.1. Regression Analysis: Overall Score vs Accuracy**

**.2. Regression Analysis: Overall Score vs Safety**

**.3. Regression Analysis: Overall Score vs Helpfulness**

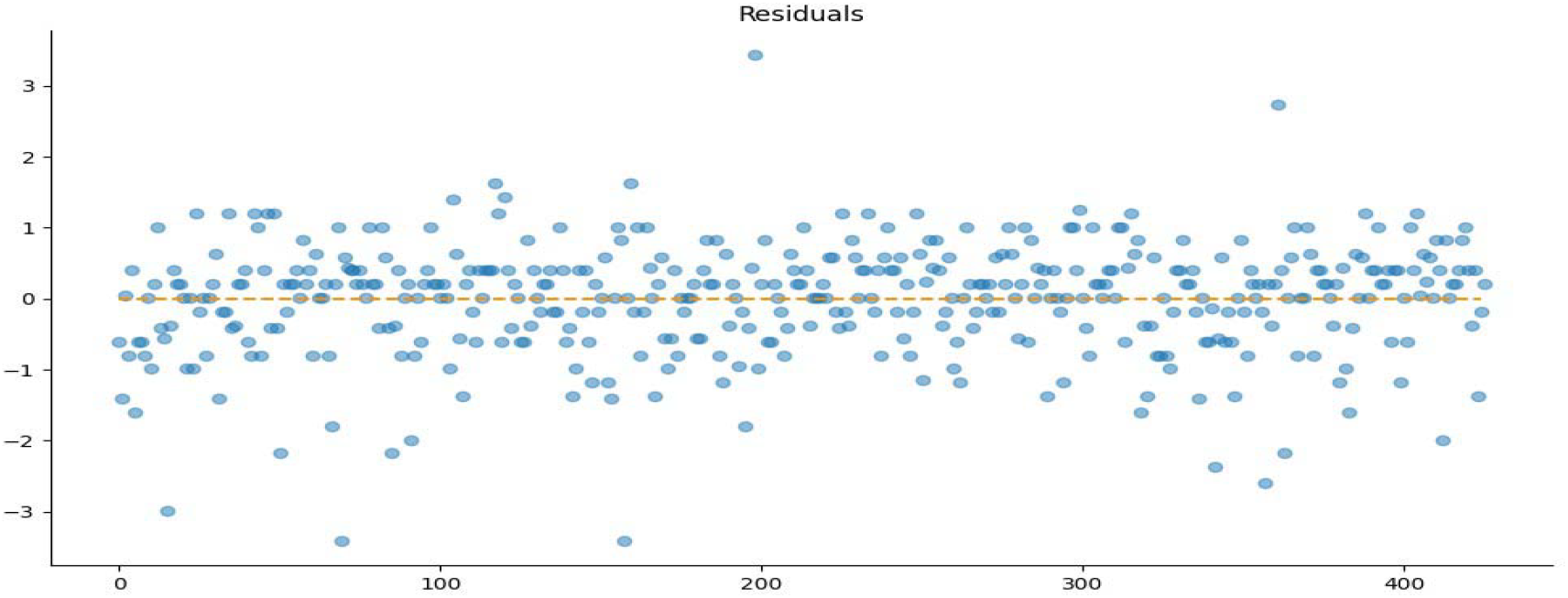

Test for Homoscedasticity: Linear Regression of Overall Score vs Helpfulness

## Appendix C.

## References

Project Genie Clinician Evaluation Group (April, 2024) https://bit.ly/clinician-evaluators-project-genie

Real statistics resource pack. Retrieved March 19, 2024. URL https://real-statistics.com/freedownload/real-statistics-resourcepack/.

Mohammad Abu-Jeyyab, Sallam Alrosan, and Ibraheem Alkhawaldeh. Harnessing large language models in medical research and scientific writing: A closer look to the future: Llms in medical research and scientific writing. High Yield Medical Reviews, 1(2), 2023.

Yasmina Al Ghadban, Huiqi Yvonne Lu, Uday Adavi, Ankita Sharma, Sridevi Gara, Neelanjana Das, Bhaskar Kumar, Renu John, Praveen Devarsetty, and Jane E Hirst. Transforming healthcare education: Harnessing large language models for frontline health worker capacity building using retrieval-augmented generation. medRxiv, pages 2023–12, 2023.

A Bosselut et al. Meditron: Open medical foundation models adapted for clinical practice. 10.21203/rs.3.rs-4139743/v1, 2024.

T.B. Brown, B. Mann, N. Ryder, M. Subbiah, J. Kaplan, P. Dhariwal, A. Neelakantan, P. Shyam, G. Sastry, A. Askell, et al. Language models are few-shot learners. ArXiv, abs/2005.14165, 2020.

Wei-Lin Chiang, Lianmin Zheng, Ying Sheng, Anastasios Nikolas Angelopoulos, Tianle Li, Dacheng Li, Hao Zhang, Banghua Zhu, Michael Jordan, Joseph E. Gonzalez, and Ion Stoica. Chatbot arena: An open platform for evaluating llms by human preference, 2024.

World Health Organization Regional Office for Africa. Stakeholders urged to take action to improve the distribution of doctors in ghana, November 7 2022. URL https://www.afro.who.int/countries/ghana/news/stakeholders-urged-take-action-improve-distribution-doctors-ghana.

World Health Organization Regional Office for Africa. Basic emergency care saving lives in ghana, June 26 2023. URL https://www.afro.who.int/photo-story/basic-emergency-care-saving-lives-ghana.

Agasthya Gangavarapu. Llms: A promising new tool for improving healthcare in lowresource nations. In 2023 IEEE Global Humanitarian Technology Conference (GHTC), pages 252–255. IEEE, 2023.

Rachel S Goodman, J Randall Patrinely Jr, Travis Osterman, Lee Wheless, and Douglas B Johnson. On the cusp: Considering the impact of artificial intelligence language models in healthcare. Med, 4(3):139–140, 2023.

Kilem L Gwet. Handbook of inter-rater reliability: The definitive guide to measuring the extent of agreement among raters. Advanced Analytics, LLC, 2014.

I. Jahan, M.T.R. Laskar, C. Peng, and J.X. Huang. A comprehensive evaluation of large language models on benchmark biomedical text processing tasks. Computers in Biology and Medicine, 171:108189, 2024. URL 10.1016/j.compbiomed.2024.108189.

Y Labrak et al. A zero-shot and few-shot study of instruction-finetuned large language models applied to clinical and biomedical tasks. ArXiv, abs/2307.12114, 2023.

I Li et al. Unleashing the power of language models in clinical settings: A trailblazing evaluation unveiling novel test design. medRxiv, 2023.

H Liu et al. Large language models are few-shot health learners. ArXiv, abs/2305.15525, 2023a.

Z Liu et al. Evaluating large language models for radiology natural language processing. ArXiv, abs/2307.13693, 2023b.

N Mehandru et al. Large language models as agents in the clinic. ArXiv, abs/2309.10895, 2023.

Paulina Boadiwaa Mensah, Nana Serwaa Quao, Sesinam Dagadu, and Project Genie Clinician Evaluation Group. All you need is context: Clinician evaluations of various iterations of a large language model-based first aid decision support tool in ghana. medRxiv, 2024. doi: 10.1101/2024.04.03.24305276.

Fred Mullan and John H. Bryant. Doctors—barefoot and otherwise: The world health organization, the united states, and global primary medical care. JAMA, 252(22):3146–3148, 1984. doi: 10.1001/jama.1984.03350220052030.

Harsha Nori, Yin Tat Lee, Sheng Zhang, Dean Carignan, Richard Edgar, Nicolo Fusi, Nicholas King, Jonathan Larson, Yuanzhi Li, Weishung Liu, et al. Can generalist foundation models outcompete special-purpose tuning? case study in medicine. arXiv preprint 2311.16452, 2023.

Python Language Reference, version 3.11. Python Software Foundation. URL http://www.python.org.

R. Rampin and V. Rampin. Taguette: open-source qualitative data analysis. Journal of Open Source Software, 6(68):3522, 2021. URL 10.21105/joss.03522.

S. Reddy, W. Rogers, V.P. Makinen, E. Coiera, P. Brown, M. Wenzel, E. Weicken, S. Ansari,P. Mathur, A. Casey, and B. Kelly. Evaluation framework to guide implementation of ai systems into healthcare settings. BMJ Health Care Inform., 28(1):e100444, 2021. URL 10.1136/bmjhci-2021-100444.

Malik Sallam. Chatgpt utility in healthcare education, research, and practice: systematic review on the promising perspectives and valid concerns. In Healthcare, volume 11, page 887. MDPI, 2023.

K. Singhal, S. Azizi, T. Tu, et al. Large language models encode clinical knowledge. Nature, 620:172–180, 2023. URL 10.1038/s41586-023-06291-2.

L. Tang, Z. Sun, B. Idnay, J.G. Nestor, A. Soroush, P.A. Elias, Z. Xu, Y. Ding,G. Durrett, J. Rousseau, C. Weng, and Y. Peng. Evaluating large language models on medical evidence summarization. medRxiv, 2023.04.22.23288967, 2023. URL 10.1101/2023.04.22.23288967.

O.R. Tanno, D. Barrett, A. Sellergren, et al. Consensus, dissensus and synergy between clinicians and specialist foundation models in radiology report generation. Research Square, 3(rs-3940387), 2024. URL 10.21203/rs.3.rs-3940387/v1.

Satvik Tripathi, Rithvik Sukumaran, and Tessa S Cook. Efficient healthcare with large language models: optimizing clinical workflow and enhancing patient care. Journal of the American Medical Informatics Association, page ocad258, 2024.

D. Van Veen, C. Van Uden, L. Blankemeier, et al. Adapted large language models can outperform medical experts in clinical text summarization. Nat Med, 20(4):543–545, 2024. URL 10.1038/s41591-024-02855-5.

Y Wang et al. Are large language models ready for healthcare? a comparative study on clinical language understanding. ArXiv, abs/2304.05368, 2023.

J. Wei, X. Wang, D. Schuurmans, M. Bosma, F. Xia, E. Chi, et al. Chain-of-thought prompting elicits reasoning in large language models. Advances in neural information processing systems, 35:24824–24837, 2022.

T. Wilhelm, J. Roos, and R. Kaczmarczyk. Large language models for therapy recommendations across 3 clinical specialties: Comparative study. J Med Internet Res, 25(10), 2023. URL https://www.jmir.org/2023/10/e49324.

Cyril Zakka, Rohan Shad, Akash Chaurasia, Alex R Dalal, Jennifer L Kim, Michael Moor, Robyn Fong, Curran Phillips, Kevin Alexander, Euan Ashley, et al. Almanac—retrievalaugmented language models for clinical medicine. NEJM AI, 1(2):AIoa2300068, 2024.

Hongjian Zhou, Boyang Gu, Xinyu Zou, Yiru Li, Sam S Chen, Peilin Zhou, Junling Liu, Yining Hua, Chengfeng Mao, Xian Wu, et al. A survey of large language models in medicine: Progress, application, and challenge. arXiv preprint 2311.05112, 2023.

